# Blood Interleukin-6 (IL-6) mRNA Expression in Alzheimer’s Disease: A Preliminary Analysis

**DOI:** 10.1101/2025.07.13.25331470

**Authors:** Mohamed Essam Hashim

## Abstract

**Introduction:** Alzheimer’s Disease (AD) is a neurodegenerative disorder characterized by amyloid protein deposition, phosphorylated tau and neurodegeneration. Interleukin-6 (IL-6) is a pro-inflammatory cytokine implicated in neurodegenerative diseases (9,10).

**Objective:** This study aimed to investigate the expression level of the IL-6 probe in blood samples from the GSE63060 dataset to determine if it significantly differs between individuals with AD, Mild Cognitive Impairment (MCI), and healthy controls (CTL), while accounting for potential confounding effects of age, gender and ethnicity.

**Methods:** Blood transcriptome data (GSE63060) from 329 participants (145 AD, 80 MCI, 104 CTL) was obtained from the Gene Expression Omnibus (GEO). Metadata, including diagnosis, age, ethnicity, and gender, was processed. The specific probe data for IL-6 ILMN_1699651 was extracted, transposed, and merged with the metadata in SPSS Statistics v25. A General Linear Model (GLM) - Univariate was employed to test the effect of Diagnosis on IL-6 expression, with Age and Gender included as covariates.

**Results:** The GLM analysis revealed **no statistically significant difference** in blood IL-6 expression among the diagnostic groups (F (2, 312) = 0.136, p=0.873).

**Conclusion:** Based on this analysis of the GSE63060 dataset, blood IL-6 mRNA expression, as measured by this probe and platform, did not significantly differ across diagnostic groups AD, MCI, and healthy control individuals when controlling for ethnicity, age, and gender. This finding suggests a need to measure plasma IL-6 protein levels, analyze other cohorts, and determine whether blood mRNA is a suitable biomarker for AD.

## 1. Introduction

Dementia and Alzheimer’s disease (AD) (the main cause of dementia) are estimated by Alzheimer’s Disease International to triple in incidence by 2050 (1). This prevalence is increasingly worrisome as the reported survival time for dementia was 3-4 years in almost 60,000 individuals (2). Although AD was initially defined clinically, where patients suffer a substantial progressive cognitive impairment affecting several domains causing functional impact on daily life, efforts developed by institutes like the International Working Group led to subsequent incorporation of biomarkers to shift AD from a clinical diagnosis to a purely biological one (1). The pathological hallmarks of AD can therefore be grouped by the presence of amyloid β protein, phosphorylated tau, and subsequent neurodegeneration (3). Increasingly important, however, are the effect of age and genetics as risk factors to AD. Although it is important to note that many genetic factors play a role in the association with the disease, it has been shown that carrying at least one allele of the APOE ε4 is a strong risk factor for developing AD (4,5). The exact causes of the disease are not fully understood and are rooted behind a complex etiology, however recently it has been shown that neuroinflammation plays a significant role in AD progression as well (6).

A meta-analysis of longitudinal and cross-sectional studies investigated the roles of inflammatory markers and their associations with mild cognitive impairment (MCI) and AD, reporting that the levels of common peripheral inflammatory cytokines, including interleukins and Tumor Necrosis Factor (TNF) were higher when associated with MCI and AD compared to control groups (7).

This was particularly significant in the case of the cytokine interleukin-6 (IL6), where it was found to be highly increased in patients with AD compared to those with MCI and controls (7). This finding may be due to the role IL-6 plays in neuroinflammation and blood brain barrier disruption by activating the JAK-STAT signaling pathway (6–8). IL-6 also promotes microglial activation resulting in phosphorylation of tau proteins in neurons (6), which is a pathological hallmark of AD.

Interestingly, collective analyses of older adults with high baseline IL-6 levels were 1.42 times more likely to develop cognitive decline in the succeeding years (9). Prolonged exposure of the brain to IL-6, despite its critical role in CNS neurodevelopmental functions, are associated with multiple neuropathological effects including AD (9,10). It is important, however, to recognize the implications of IL-6 mediated pathogenesis in other pathologic states including infection, autoimmunity and trauma, where blocking of IL-6 trans-signaling at a cellular or molecular level can to lead to improvements in the negative effects IL-6 has on microglia and astrocytosis in mice (8). Additionally, mutations in IL-6 may potentiate the risk of developing AD, as increased activation of IL-6 leads to neurodegeneration (11). Such effects of inflammatory cytokines including TNF-alpha, IL-1 and IL-6 demonstrate the importance of inflammation in neurodegenerative disorders and how we can use these molecules to detect and possibly treat neuroinflammation in AD (11).

Over the last two decades, AD detection has shifted from an imperfect clinical-only diagnosis to more advanced techniques ranging from amyloid and tau positron emission tomography (PET) to cerebrospinal fluid analyses showing high sensitivity and specificity in predicting AD conversion from MCI, in one study (12,13). Despite these validated findings, plasma-based biomarker analyses are gaining popularity due to the expensive and limited availability of PET, as well as the invasive nature of CSF collection both of which limit the potential for large-scale screening (13). Gene expression studies for circulating levels of risk factors in AD, can therefore facilitate the process of diagnosing AD by detecting biomarker level RNA extraction and analysis, which can be done from tissue samples including blood (11,14). This allows for early-detection of AD and neurodegeneration without exposing patients to invasive procedures and costly imaging, therefore providing new pathways for disease diagnosis and treatment. Despite the promising nature of gene expression studies (transcriptomics), they do not come without limitations and challenges in studying them. RNA quality and tissue profiles, whether the sample derives from brain tissue or blood, can be variable and hard to maintain (14). While peripheral samples of genes encoding inflammatory cytokines and transcriptomic studies would be promising markers for AD detection, they may not fully reflect brain changes as CNS amyloid deposition (14). This warrants the need for further studies involving blood transcriptomics and their significance in AD correlation and diagnosis.

One study conducted by the National Center for Biotechnology Information Gene Expression Omnibus (GEO) to identify AD biomarkers by blood, which is a large AddNeuroMed AD cohort, provided their dataset publicly with blood source RNA for AD gene biomarkers and demographic characteristics (15). One subseries of this cohort is the batch 1 GSE63060 dataset where we have extracted the IL-6 probe available in the dataset to analyze whether there is a significant association between IL-6 peripheral expression levels and diagnostic group (AD, MCI, Controls) controlling for age and gender.

Given the role IL-6 plays in neuroinflammation and its potential relevance to AD, this study aims to investigate the levels of IL-6 probes in patients with AD, MCI and healthy controls after accounting for the potential influence of age and gender.

## 2. Methods

### 2.1 Data Acquisition

The dataset GSE63060 was acquired from the NCBI Gene Expression Omnibus (GEO) (http://www.ncbi.nlm.nih.gov/geo/query/acc.cgi?acc=GSE63063) on May 7, 2025. (15)

The data was generated using the Illumina HumanHT-12 V3.0 expression beadchip platform GEO accession: GPL6947; http://www.ncbi.nlm.nih.gov/geo/query/acc.cgi?acc=GPL6947). (16)

The **GSE63060_series_matrix.txt.gz** file was downloaded, containing the processed and normalized gene expression matrix (note that in the supplementary file section on the GSE63060 GEO viewer the normalized tab delimited text file can be found).

Text file (**GSE63060_series_matrix.txt.gz**) was then imported into Microsoft Excel for data cleaning and preparation.

### 2.2. Data Cleaning and Preparation

Initial data inspection was performed using Microsoft Excel v16.78.3.

The metadata for each sample, including status (AD, MCI, control [CTL]), Age, Gender, and Ethnicity, was extracted and organized into a separate sheet within the GSE63063_Series_Matrix Excel workbook. This was done by manually locating metadata header lines (first column; prefixed with an exclamation mark ‘!’, e.g., !Sample_title), and copying and pasting into a new sheet within the workbook by ‘Paste Special’ then transpose to transpose the rows and columns for saving and importing to SPSS. After which the headers that contain the same values for each sample including !Sample_contact_name and !Sample_type where removed only leaving the Sample Geo accession (which was renamed to SampleID) and demographic characteristics (Age, Gender, Ethnicity). The metadata headings were renamed to their respective variable names (SampleID, Age, Gender, Ethnicity).

This cleaned metadata was saved as a comma-separated values (CSV) file GSE63060_Metadata_Cleaned.csv.

The specific Probe ID corresponding to the IL-6 gene on the GPL6947 platform was identified by searching the platform’s annotation file GPL6947.annot, downloaded from the GPL6947 GEO page. The identified Probe ID for IL-6 was ILMN_1699651.

The expression data matrix (starting with ID_REF and GSM sample IDs as headers, tabdelimited) was prepared by saving a TXT file (GSE63060_Expression_RawMatrix.txt) after removing headers by copying and pasting the expression data into another new sheet within the GSE63063_Series_Matrix workbook. Preparation of IL-6 Expression Data: Due to the analysis focus on a single probe, the expression dataset (Probes as rows, Samples as columns) in Excel was filtered to select only the row corresponding to the IL-6 Probe ID ILMN_1699651 with GSM sample IDs. The resulting single-row dataset contained the IL-6 expression values across all samples. This single-row IL-6 dataset was then transposed in Excel by copy and ‘Paste Special’ with ‘Transpose’. Then was exported as GSE63060_Expression_RawMatrix.txt

Data analysis was conducted using IBM SPSS Statistics v25.

The cleaned metadata file GSE63060_Metadata_Cleaned.csv was imported into SPSS using the Text Import Wizard, ensuring variables were correctly defined (SampleID as String/Nominal, Diagnosis/Gender/Ethnicity as String/Nominal or Numeric/Nominal with value labels, Age as Numeric/Scale)

The expression data file GSE63060_Expression_RawMatrix.txt was imported into SPSS using the Text Import Wizard. Key wizard settings included ‘Yes’ for variable names on the first line, data starting on line 3, ‘Tab’ delimiter, ID_REF as String/Nominal, GSM columns as Numeric/Scale.

The transposed dataset containing IL-6 expression was imported into SPSS using the Text Import Wizard. Key wizard settings included ‘Yes’ for variable names on the first line, data starting on line 2, Tab delimiter, ID_Ref as String/Nominal, ILMN_1699651 as Numeric/Scale.

The transposed dataset imported into SPSS was cleaned by renaming the ID_Ref variable to SampleID and the IL-6 expression variable to IL6_Expression.

Finally, the transposed IL-6 expression dataset was merged with the cleaned metadata dataset in SPSS using SampleID as the key variable. SPSS steps: Data > Merge Files > Add Variables, selecting the transposed IL-6 dataset and matching cases on SampleID. The resulting dataset contained all sample metadata and the IL6_Expression variable, with each row representing a unique sample.

### 2.3. Statistical Analysis

Descriptive statistics, including frequencies for categorical variables (Diagnosis, Gender, Ethnicity) and means and standard deviations for continuous variables (Age, IL6_Expression), were computed using the SPSS Frequencies and Descriptives procedures (Analyze > Descriptive Statistics > Frequencies) and (Analyze > Descriptive Statistics > Descriptives). Means of IL-6 expression within each diagnostic group were also calculated using the SPSS Compare Means > Means procedure.

To assess whether mean blood IL-6 expression differed significantly between diagnostic groups (AD, MCI, CTL) while controlling for the potential confounding effects of age and gender, a General Linear Model (GLM) - Univariate analysis was conducted using the SPSS Analyze > General Linear Model > Univariate procedure.

In the GLM, IL-6 expression (IL6_Expression) was set as the Dependent Variable.

Diagnosis (AD, MCI, CTL) and Gender were included as Fixed Factors.

Age was included as a Covariate.

The model included the default Full factorial.

Estimated marginal means for Diagnosis, Homogeneity tests (Levene’s), Estimates of effect size, Observed power, Parameter estimates were requested in the Options dialog.

Post-hoc tests for Diagnosis were not interpreted due to the non-significant overall effect of Diagnosis in the GLM.

Statistical significance was determined using an alpha level of α=0.05.

## 3. Results

### 3.1. Participant Characteristics

The final analysis included data from 329 participants (145AD, 80 MCI, 104 CTL).

The demographic characteristics of the cohort, stratified by diagnostic group, are presented in Table 1.

**Table 1.** Participant Demographics by Diagnostic Group.

| Diagnosis |  |  |  |  |  |
| --- | --- | --- | --- | --- | --- |
|  |  | Frequency | Percent | Valid Percent | Cumulative Percent |
| Valid | AD | 145 | 44.1 | 44.1 | 44.1 |
|  | CTL | 104 | 31.6 | 31.6 | 75.7 |
|  | MCI | 80 | 24.3 | 24.3 | 100.0 |
|  | Total | 329 | 100.0 | 100.0 |  |
| Gender |  |  |  |  |  |
|  |  | Frequency | Percent | Valid Percent | Cumulative Percent |
| Valid | Female | 200 | 60.8 | 60.8 | 60.8 |
|  | Male | 129 | 39.2 | 39.2 | 100.0 |
|  | Total | 329 | 100.0 | 100.0 |  |
| Ethnicity |  |  |  |  |  |
|  |  | Frequency | Percent | Valid Percent | Cumulative Percent |
| Valid | Other Caucasian | 42 | 12.8 | 12.8 | 12.8 |
|  | Unknown | 5 | 1.5 | 1.5 | 14.3 |
|  | Western European | 282 | 85.7 | 85.7 | 100.0 |
|  | Total | 329 | 100.0 | 100.0 |  |

### 3.2. IL-6 Expression Levels

Mean IL-6 expression levels were calculated for each diagnostic group. Descriptive statistics for IL-6 expression are presented in Table 2.

**Table 2.** IL-6 Expression (Mean ± SD) by Diagnostic Group.

| Report |  |  |  |
| --- | --- | --- | --- |
| IL6_Expression |  |  |  |
| Diagnosis | Mean | N | Std. Deviation |
| AD | 7.4927 | 145 | .07038 |
| CTL | 7.5018 | 104 | .06102 |
| MCI | 7.4968 | 80 | .06892 |
| Total | 7.4966 | 329 | .06711 |

**Table 3.** Tests of Between-Subjects Effects for IL-6 Expression.

| Tests of Between-Subjects Effects |  |  |  |  |  |  |  |  |
| --- | --- | --- | --- | --- | --- | --- | --- | --- |
| Dependent Variable: IL6_Expression |  |  |  |  |  |  |  |  |
| Source | Type III Sum of Squares | df | Mean Square | F | Sig. | Partial Eta Squared | Noncent. Parameter | Observed Power<sup>b</sup> |
| Corrected Model | .061<sup>a</sup> | 16 | .004 | .836 | .644 | .041 | 13.381 | .575 |
| Intercept | 130.933 | 1 | 130.933 | 28843.341 | .000 | .989 | 28843.341 | 1.000 |
| Age | .011 | 1 | .011 | 2.506 | .114 | .008 | 2.506 | .352 |
| Diagnosis | .001 | 2 | .001 | .136 | .873 | .001 | .272 | .071 |
| Gender | .015 | 1 | .015 | 3.318 | .069 | .011 | 3.318 | .443 |
| Ethnicity | .003 | 2 | .002 | .336 | .715 | .002 | .672 | .104 |
| Diagnosis * Gender | .006 | 2 | .003 | .650 | .523 | .004 | 1.299 | .159 |
| Diagnosis * Ethnicity | .005 | 3 | .002 | .337 | .798 | .003 | 1.012 | .116 |
| Gender * Ethnicity | .019 | 2 | .009 | 2.052 | .130 | .013 | 4.103 | .421 |
| Diagnosis * Gender * Ethnicity | .014 | 3 | .005 | 1.037 | .376 | .010 | 3.111 | .281 |
| Error | 1.416 | 312 | .005 |  |  |  |  |  |
| Total | 18490.907 | 329 |  |  |  |  |  |  |
| Corrected Total | 1.477 | 328 |  |  |  |  |  |  |
| a R Squared = .041 (Adjusted R Squared = -.008) |  |  |  |  |  |  |  |  |
| b Computed using alpha = |  |  |  |  |  |  |  |  |

Visual inspection of IL-6 expression distribution across groups is shown in Figure 1.

**Figure 1.**
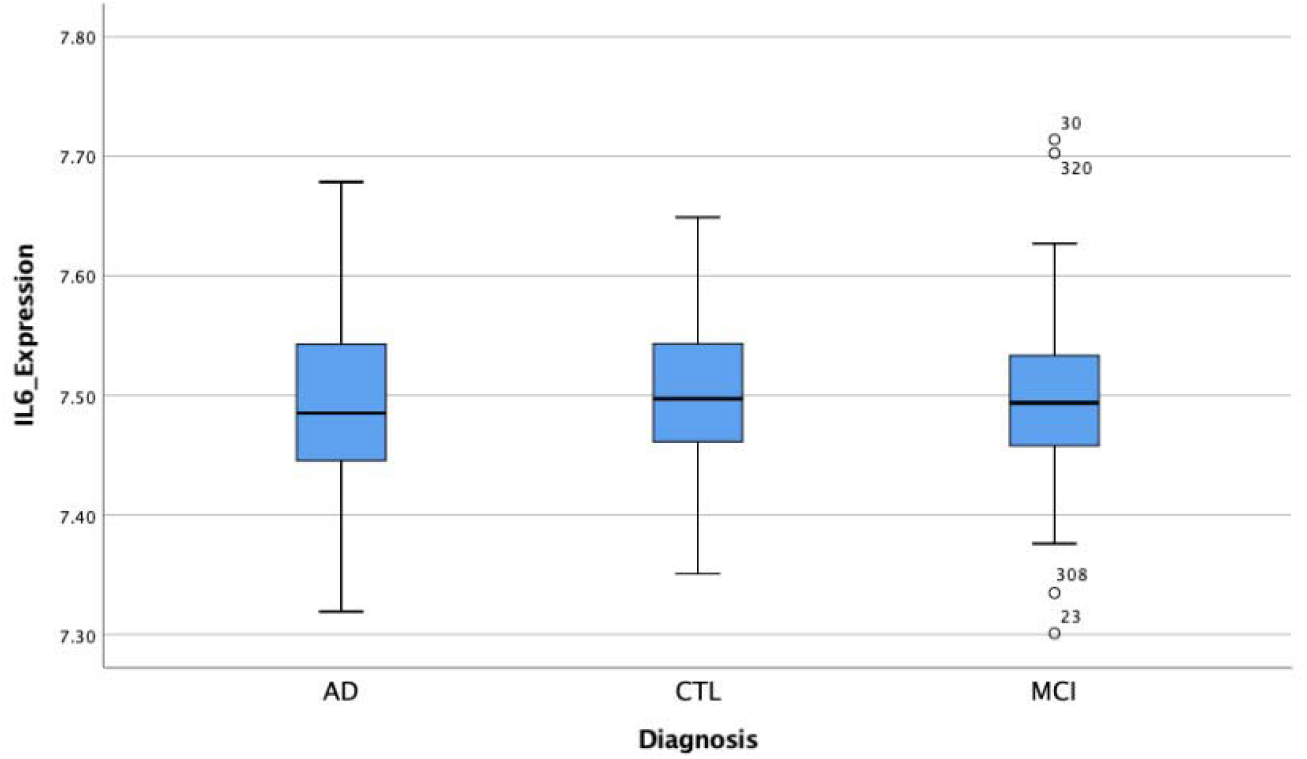
Box plot of IL-6 expression levels by diagnostic group in the GSE63060 dataset. Boxes represent the interquartile range, the line inside the box is the median.

### 3.3. General Linear Model Analysis for IL-6 Expression

Levene’s Test for Equality of Error Variances was conducted to assess the assumption of homogeneity of variances for IL-6 expression across diagnostic groups. The test was not statistically significant, based on p-value (p=.148). (Note: Comment on implications in Discussion if significant).

The GLM analysis assessing the effect of Diagnosis on IL-6 expression, controlling for Age and Gender, revealed no statistically significant main effect of Diagnosis (F (2, 312) =.136, p=0.873). This result indicates that, after accounting for age and gender, there is no statistically significant evidence in this dataset to conclude that the mean blood IL-6 expression differs among the AD, MCI, and CTL groups.

The estimated marginal means for IL-6 expression, adjusted for age and gender, were 7.494 for the AD group, 7.484 for the MCI group, and 7.490 for the CTL group.

Post-hoc pairwise comparisons for Diagnosis were not interpreted as the overall main effect of Diagnosis was not statistically significant (p>0.05).

## 4. Discussion

This study investigated blood IL-6 mRNA expression in the GSE63060 dataset and found no statistically significant difference in expression levels between individuals with AD, MCI, and healthy controls when controlling for participant age and gender.

Interleukin-6 is a well-established pro-inflammatory cytokine (8–10). Given the significant role of neuroinflammation in AD pathogenesis (6,10), this study initially hypothesized that IL-6 expression in peripheral blood might be altered in AD patients. However, our finding does not support this hypothesis for blood mRNA levels in this specific cohort. This contrasts with findings from other studies that reported elevated IL-6 protein levels in the blood or cerebrospinal fluid of AD patients (9,10). It is important to note that mRNA levels and protein levels do not always perfectly correlate due to complex post-transcriptional and posttranslational regulatory mechanisms (14). This lack of association between IL-6 and AD diagnosis could potentially be due to differences in cohort characteristics, disease stage, or specific methodologies used across studies (7,10).

Furthermore, although genetic variants in the IL6 gene have been linked to AD risk (11), suggesting involvement of IL-6 pathways, our analysis of blood expression in this dataset did not reveal a significant difference tied to diagnosis. The initial hypothesis was biologically grounded given IL-6’s role in inflammation relevant to AD. However, our data suggest that blood mRNA expression of IL-6, as measured by the GPL6947 platform in the GSE63060 cohort, may not be a sufficiently sensitive or reliable marker to differentiate diagnostic groups. This non-significant finding contributes valuable information by highlighting potential limitations of using blood IL-6 mRNA alone as a diagnostic biomarker in this context.

Several limitations should be considered. First, this analysis is based on a single dataset (GSE63060) and the findings may be specific to this cohort. Second, we focused on a single probe representing IL-6 mRNA; analysis of other IL-6 related transcripts or using different gene expression measurement technologies could yield different results. Third, blood-based markers may not fully capture the inflammatory state within the central nervous system to the same extent as measures from CSF would. Finally, our analysis focused on mRNA expression; the levels and activity of IL-6 protein may be more directly relevant to its biological function and could show different patterns.

Future studies should investigate blood IL-6 protein levels in the GSE63060 cohort if corresponding samples are available, or in independent AD cohorts, including the second batch of AddNeuroMed data (GSE63061), to determine if protein levels show a different relationship with diagnosis. Further research could also explore IL-6 expression within specific immune cell populations in blood (e.g., T-lymphocytes), or investigate the combined potential of IL-6 with other candidate blood biomarkers for AD (APOE ε4, for instance).

## 5. Conclusion

In conclusion, this analysis of the GSE63060 dataset found no statistically significant difference in blood IL-6 mRNA expression levels between individuals with Alzheimer’s Disease, Mild Cognitive Impairment, and healthy controls, after accounting for age and gender. While biologically plausible, blood IL-6 mRNA expression did not serve as a significant diagnostic marker in this study, suggesting the need for further investigation of IL-6 using alternative measurements (protein levels, CSF) or in different cohorts.

## Data Availability

All data analyzed in this study are publicly available from the NCBI Gene Expression Omnibus (GEO) under accession number GSE63060. No new data were generated. https://www.ncbi.nlm.nih.gov/geo/query/acc.cgi?acc=GPL6947

https://ftp.ncbi.nlm.nih.gov/geo/series/GSE63nnn/GSE63060/matrix/GSE63060_series_matrix.txt.gz

https://www.ncbi.nlm.nih.gov/geo/query/acc.cgi?acc=GPL6947

## References

1. Scheltens P, Strooper BD, Kivipelto M, Holstege H, Chételat G, Teunissen CE, et al. Alzheimer’s disease. Lancet Lond Engl. 2021 Apr 24;397(10284):1577–90.

2. Mayeda ER, Glymour MM, Quesenberry CP, Johnson JK, Pérez-Stable EJ, Whitmer RA. Survival After Dementia Diagnosis in Five Racial/Ethnic Groups. Alzheimers Dement J Alzheimers Assoc. 2017 Jul;13(7):761–9.

3. Jack CR, Bennett DA, Blennow K, Carrillo MC, Dunn B, Haeberlein SB, et al. NIA-AA Research Framework: Toward a biological definition of Alzheimer’s disease. Alzheimers Dement J Alzheimers Assoc. 2018 Apr;14(4):535–62.

4. The effect of APOE and other common genetic variants on the onset of Alzheimer’s disease and dementia: a community-based cohort study - The Lancet Neurology [Internet]. [cited 2025 May 7]. Available from: https://www.thelancet.com/journals/laneur/article/PIIS14744422(18)30053-X/abstract

5. Ng TKS, Beck T, Boyle P, Dhana K, Desai P, Evans DA, et al. APOE4, Blood Neurodegenerative Biomarkers, and Cognitive Decline in Community-Dwelling Older Adults. JAMA Netw Open. 2025 May 7;8(5):e258903.

6. Khan AW, Farooq M, Hwang MJ, Haseeb M, Choi S. Autoimmune Neuroinflammatory Diseases: Role of Interleukins. Int J Mol Sci. 2023 Apr 27;24(9):7960.

7. Leonardo S, Fregni F. Association of inflammation and cognition in the elderly: A systematic review and meta-analysis. Front Aging Neurosci [Internet]. 2023 Feb 6 [cited 2025 May 7];15. Available from: https://www.frontiersin.org/journals/agingneuroscience/articles/10.3389/fnagi.2023.1069439/full

8. Campbell IL, Erta M, Lim SL, Frausto R, May U, Rose-John S, et al. Trans-Signaling Is a Dominant Mechanism for the Pathogenic Actions of Interleukin-6 in the Brain. J Neurosci. 2014 Feb 12;34(7):2503–13.

9. Bradburn S, Sarginson J, Murgatroyd CA. Association of Peripheral Interleukin-6 with Global Cognitive Decline in Non-demented Adults: A Meta-Analysis of Prospective Studies. Front Aging Neurosci. 2018 Jan 8;9:438.

10. Wu YY, Hsu JL, Wang HC, Wu SJ, Hong CJ, Cheng IHJ. Alterations of the Neuroinflammatory Markers IL-6 and TRAIL in Alzheimer’s Disease. Dement Geriatr Cogn Disord EXTRA. 2015 Nov 24;5(3):424–34.

11. Dhapola R, Hota SS, Sarma P, Bhattacharyya A, Medhi B, Reddy DH. Recent advances in molecular pathways and therapeutic implications targeting neuroinflammation for Alzheimer’s disease. Inflammopharmacology. 2021 Dec;29(6):1669–81.

12. Santangelo R, Masserini F, Agosta F, Sala A, Caminiti SP, Cecchetti G, et al. CSF ptau/Aβ42 ratio and brain FDG-PET may reliably detect MCI “imminent” converters to AD. Eur J Nucl Med Mol Imaging. 2020 Dec 1;47(13):3152–64.

13. Muir RT, Ismail Z, Black SE, Smith EE. Comparative methods for quantifying plasma biomarkers in Alzheimer’s disease: Implications for the next frontier in cerebral amyloid angiopathy diagnostics. Alzheimers Dement. 2023 Oct 31;20(2):1436–58.

14. Bagyinszky E, Giau VV, An SA. Transcriptomics in Alzheimer’s Disease: Aspects and Challenges. Int J Mol Sci. 2020 May 15;21(10):3517.

15. GEO Accession viewer [Internet]. [cited 2025 May 7]. Available from: https://www.ncbi.nlm.nih.gov/geo/query/acc.cgi?acc=GSE63060

16. Illumina HumanHT-12 V3.0 expression beadchip [Internet]. [cited 2025 May 7]. Available from: https://www.ncbi.nlm.nih.gov/geo/query/acc.cgi?acc=GPL6947

